# Outbreak.info genomic reports: scalable and dynamic surveillance of SARS-CoV-2 variants and mutations

**DOI:** 10.1101/2022.01.27.22269965

**Authors:** Karthik Gangavarapu, Alaa Abdel Latif, Julia L. Mullen, Manar Alkuzweny, Emory Hufbauer, Ginger Tsueng, Emily Haag, Mark Zeller, Christine M. Aceves, Karina Zaiets, Marco Cano, Jerry Zhou, Zhongchao Qian, Rachel Sattler, Nathaniel L Matteson, Joshua I. Levy, Raphael TC Lee, Lucas Freitas, Sebastian Maurer-Stroh, GISAID core and curation team, Marc A. Suchard, Chunlei Wu, Andrew I. Su, Kristian G. Andersen, Laura D. Hughes

## Abstract

The emergence of SARS-CoV-2 variants of concern has prompted the need for near real-time genomic surveillance to inform public health interventions. In response to this need, the global scientific community, through unprecedented effort, has sequenced and shared over 10 million genomes through GISAID, as of May 2022. This extraordinarily high sampling rate provides a unique opportunity to track the evolution of the virus in near real-time. Here, we present outbreak.info, a platform that currently tracks over 40 million combinations of PANGO lineages and individual mutations, across over 7,000 locations, to provide insights for researchers, public health officials, and the general public. We describe the interpretable and opinionated visualizations in the variant and location focussed reports available in our web application, the pipelines that enable the scalable ingestion of heterogeneous sources of SARS-CoV-2 variant data, and the server infrastructure that enables widespread data dissemination via a high performance API that can be accessed using an R package. We present a case study that illustrates how outbreak.info can be used for genomic surveillance and as a hypothesis generation tool to understand the ongoing pandemic at varying geographic and temporal scales. With an emphasis on scalability, interactivity, interpretability, and reusability, outbreak.info provides a template to enable genomic surveillance at a global and localized scale.

## Introduction

In December 2019, a series of cases of pneumonia of unknown origin appeared in Wuhan, China, and on 7 January 2020, the virus responsible for the diseases was identified as a novel coronavirus, SARS-CoV-2^1^. The first SARS-CoV-2 genome was made publicly available on 10 January 2020^2^. Since then, the global scientific community, through an unprecedented effort, has sequenced and shared over 10 million genomes through GISAID, as of May 2022^3,4^. To keep track of the evolving genetic diversity of SARS-CoV-2, Rambaut *et al*. developed a dynamic phylogeny-informed nomenclature (PANGO) to classify SARS-CoV-2 lineages^5^. As of May 2022, over 2,000 lineages have been designated, which has enabled public health agencies such as Public Health England (PHE), the Centers for Disease Control (CDC), and the World Health Organization (WHO) to identify Variants of Concern (VOC), Variants of Interest (VOI/VUI), and Variants Under Monitoring (VUM/VBM) based on the phenotypical characterization of these lineages^6^. Currently, there are five designated VOCs: B.1.1.7* (Alpha; * denotes the lineage and any of its sub lineages) lineage resulting in increased transmissibility^7^, B.1.351* (Beta) lineage exhibiting immune evasion^8^, the P.1* (Gamma) lineage exhibiting immune evasion^9^, the B.1.617.2* lineage exhibiting increased transmissibility due to the P681R mutation in the Spike gene^10^, and more recently, the B.1.1.529* (Omicron) lineage exhibiting very rapid growth and the ability to substantially avoid antibody neutralization^11,12^.

The emergence of VOCs with fitness advantages has led to global “sweeps’’ with newly emerged VOCs displacing previously circulating variants. More importantly, the growth of each VOC has led to a renewed surge in infections worldwide. This has prompted the need for near real-time genomic surveillance to inform early public health interventions to control the rise of infections. In response to this need, thousands of academic, non-academic, and public health labs have been depositing sequences predominantly on the sharing platform of the GISAID Initiative^4,13^. This extraordinarily high sampling rate of infecting viruses provides a unique opportunity to track the evolution of the virus in near real-time. For example, in December 2021 alone, over a million new genomes were submitted to GISAID^14^. Traditionally, phylodynamic approaches have been employed to retrospectively characterize lineage dynamics during outbreaks of viruses such as Zika^15–17^, West Nile^18^ and Ebola viruses^19,20^. Existing tools like NextStrain^21^ and frameworks such as Microreact^22^ primarily rely on a phylogeny to elucidate transmission chains and monitor the evolution of the virus. However, these tools were not designed to track thousands of new genomes per day, and given that building phylogenies for large sets of genomes is computationally intensive and time consuming, obtaining timely insights from the data is often problematic^23^. However, the high sampling rate of the virus has opened up the possibility of tracking the pandemic using the available near real-time genomic data without the need for computationally intensive modeling.

Here, we present outbreak.info, a platform that currently tracks over 40 million combinations of PANGO lineages and individual mutations, across over 7,000 locations, to provide insights for researchers, public health officials, and the general public. In the following sections, we describe the data pipelines that enable the scalable ingestion and standardization of heterogeneous data on SARS-CoV-2 variants, the server infrastructure that enables the dissemination of the processed data, and the client-side applications that provide intuitive visualizations of the underlying data.

## Results

The growth rate of a given viral lineage is a function of epidemiology and its intrinsic biological properties (**Fig 1a**). For example, the B.1.177 lineage, characterized by an A222V amino acid substitution in the spike gene, increased in prevalence in Europe during the summer of 2020^24^. While initially thought to be more transmissible, it was eventually shown that the increase in prevalence was due to a resurgence in travel and not due to increased transmissibility. In contrast, a few months later, the B.1.1.7 lineage was shown to be 40-60% more transmissible than previously circulating lineages and this intrinsic biological property led to the rapid growth in its prevalence worldwide^25,26^. Epidemiological factors such as mobility^27,28^, mask usage^29^, and public health interventions^30^ vary over time and across geographies worldwide, while biological properties are a function of the mutations found in a given lineage (**Fig 1a**). Hence, to maximize the utility of genomic data for surveillance, we built outbreak.info to enable the exploration of genomic data across three dimensions: geography, time, and lineages/mutations. We use the PANGO nomenclature to estimate the prevalence of SARS-CoV-2 lineages over time and at varying geographic scales. Using a phylogenetically-informed nomenclature allows us to determine genetic features such as the “characteristic mutations” of a lineage without directly building a global phylogeny. By avoiding a global phylogeny, we can update our databases daily using the continuously growing number of SARS-CoV-2 genomes. In addition, we closely track reports from health agencies such as the PHE, the CDC and the WHO that designate VOC/VOI/VUMs based on epidemiological analyses. In addition to genomic data, the server also ingests two other types of data: (1) epidemiological data curated by Johns Hopkins University^31^, and (2) public literature, clinical trial, protocol, and dataset metadata from sources such as bioRxiv, medRxiv, and LitCovid^32^. Here, we describe how each of these data sources is used in cohesion to assist in genomic surveillance.

**Figure 1.**
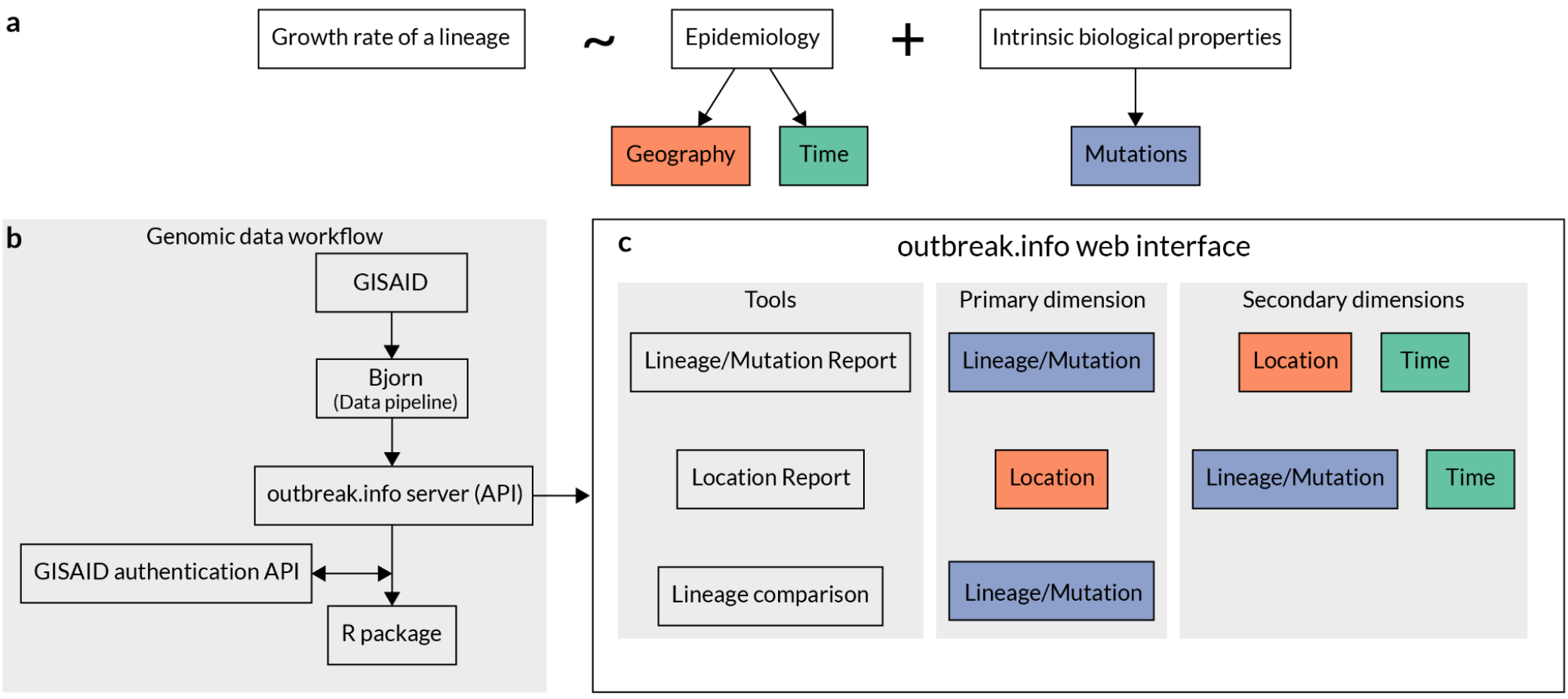
outbreak.info enables the exploration of genomic data across three dimensions. **a**, Growth rate of a lineage is a function of epidemiology and intrinsic biological properties of a lineage. Further, epidemiology varies over time and by geography while intrinsic biological properties are determined by the mutations present in a given lineage. **b**, Genomic data is ingested from GISAID, processed using the custom-built data pipeline, `Bjorn`, and stored on a server which can be accessed via an Application Programming Interface (API). The API is consumed by two clients: A JavaScript based web client and an R package that provides programmatic access by authenticating against GISAID credentials. **c**, The web interface contains three tools that allow exploration of genomic data across three different dimensions: lineage/mutation, time, and geography.

**Figure 2.**
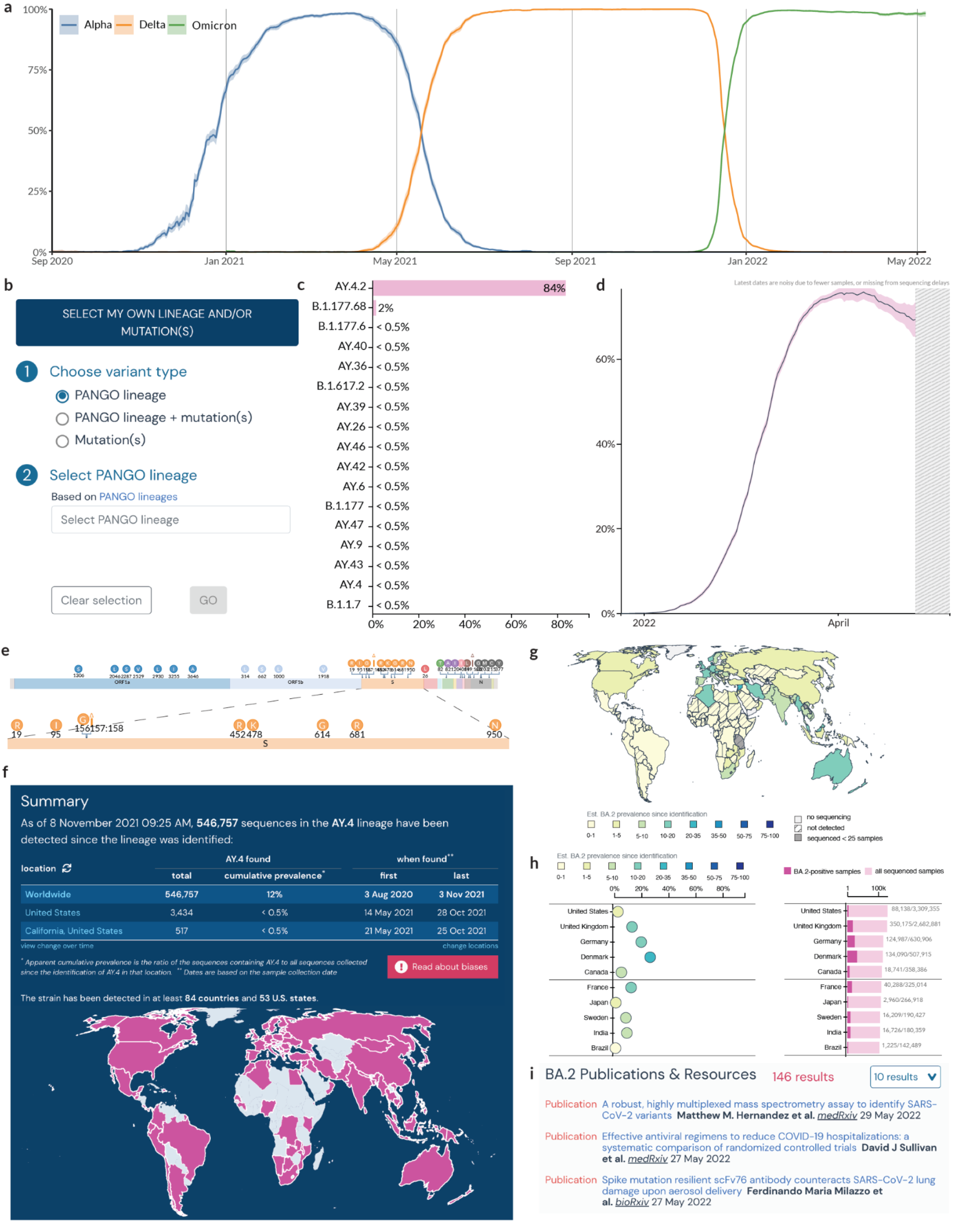
Lineage and/or Mutation Tracker. **a**, Prevalence of VOCs in the United Kingdom from Sep 2020 to May 2022. **b**, Search and filter options for Lineage/Variant of Concern tracker. **c**, Prevalence of S:Y145H + S:A222V mutations across different lineages globally. **d**, Prevalence of BA.2 in the United Kingdom. **e**, Mutation map showing the characteristic mutations of AY.4. **f**, Summary statistics of BA.2 lineage. **g**, Geographic distribution of the cumulative prevalence of BA.2 lineage globally. **h**, Cumulative prevalence of BA.2 in each country globally. **i**, Research articles, and datasets related to BA.2.

The overall workflow of genomic data is shown in **Fig 1b**. Genomic data is ingested from GISAID, processed via a custom-built data pipeline, `Bjorn`, and stored on a server which can be accessed via an application programming interface (API). We built two client-side applications, a web interface and an R package which consume this API (**Fig 1b**). The web interface consists of three main tools focussing on different facets of the underlying genomic data: (1) Lineage and/or Mutation Tracker, (2) Location Tracker, and (3) Lineage Comparison Tool. We designed an opinionated interface for each tool that focuses on one primary dimension of the genomic data with additional customizability of one or more secondary dimensions (**Fig 1c**). The Lineage and/or Mutation Tracker focus on a specific lineage, mutation or a combination of these. The Location Tracker focuses on a given location and provides a snapshot of currently circulating lineages. Finally, the Lineage Comparison Tool can be used to explore the prevalence of mutations across different lineages. In addition to the web interface, we have built an R package that authenticates against GISAID credentials and allows programmatic access to the processed data for downstream analyses.

### Lineage and/or Mutation Tracker

The ongoing SARS-CoV-2 pandemic has been punctuated by the emergence of VOCs with fitness advantages over previously circulating variants, resulting in “waves” of infections. **Fig 1a** shows the changing prevalence of the three most dominant VOCs in the United Kingdom, but this phenomenon is observed globally with heterogeneity across geography. A fundamental part of genomic surveillance is to identify the emergence of such variants by closely tracking the growth of circulating lineages. Given the geographic variation in epidemiological, social, and economic factors, it is important to estimate variant prevalence at varying geographic scales. The Lineage/Mutation Tracker can be used to dynamically query the temporal and geographic variation in the prevalence of a (i) VOC/VOI and its sublineages (e.g., Delta and its sublineages), (ii) a lineage (e.g., B.1.1.7), (iii) a lineage and one or more mutations (e.g., B.1.1.7 with S:E484K), (iv) a mutation (e.g., S:E484K), or (iv) a group of mutations (e.g., S:E484K and S:N501Y) (**Fig 1b**). In addition, users can specify various location scales, such as a country, state, or county (or their local equivalents), to estimate the prevalence of a given lineage and/or mutations. To provide meaningful insights from these prevalence estimates, we designed an opinionated interface to address a specific set of questions listed in Table 1.

**Table 1.** Questions addressed by the Lineage and/or Mutation Tracker.

| Question | Relevant visual elements |
| --- | --- |
| What is the prevalence of a set of mutations within different lineages? | Mutations such as S:N501Y, S:DEL69/70, and S:E484K have been shown to have functional impact on the phenotype exhibited by a lineage such as increased pathogenicity or immune evasion<sup>33,34</sup>. Furthermore, these mutations have been acquired independently by many lineages. Convergent evolution can be used as a metric to assess the importance of any advantage conferred on a lineage by a mutation. Hence, if a query contains a set of mutations (e.g., S:E484K and S:N501Y), we estimate the prevalence of that set of mutations across all lineages globally. (**Fig 2c**). |
| What is the trend shown by the prevalence of a lineage and/or a set of mutations over time? | Tracking the growth rate of a lineage or a set of mutations over time is very important to inform public health interventions. We estimate the prevalence of a given query as a proportion of the total number of sequences collected on a given day at a given location. To convey the uncertainty in estimating the prevalence, we calculate binomial proportion confidence intervals using Jeffrey's interval (**Fig 2d**). |
| What are the “characteristic mutations” of a lineage? | The mutations that are characteristic of a lineage can be used to generate hypotheses about the phenotype exhibited by a lineage based on prior studies on the functional impact of mutations. This is especially important to assess any potential impact a lineage might have on therapeutics such as monoclonal antibody drugs. We define the “characteristic mutations” of a lineage as those mutations found in at least 75% of the genomes classified as the lineage (**Fig 2e**). These mutations are displayed in a “mutation map”. |
| What is the total number of sequences that belong to a lineage and/or a set of mutations?<br>In how many countries was a lineage and/or a set of mutations detected?<br>When was this lineage and/or a set of mutations first detected? | In order to assess how quickly a variant spread and the extent of the geographic spread, we show summary of relevant statistics such as the total number of sequences that match the query, the cumulative prevalence of these mutations, the first and last date a sequence matching the query was detected worldwide for a customizable set of locations (**Fig 2f**). |
| What is the geographic prevalence of a lineage and/or a set of mutations? | Many lineages including VOCs Beta and Gamma show variation in growth rates across different locations. Hence, it is essential to be able to access the geographic distribution of a given lineage. To facilitate this, we show the cumulative prevalence of lineages since they were first detected across the sub-admin levels of a given location for a lineage/mutation query (**Fig 2g**). Choropleths are useful visual elements to map geographic variation in prevalence but to further highlight the uncertainty in these estimates and to account for cognitive biases in evaluating locations with different areas, we use a dot chart to show the uncertainty in the point estimate of prevalence over the last 60 days and a bar chart to show the number of sequences used |
|  | to calculate it (**Fig 2h**). These two charts can be sorted by the prevalence of the query or the total number of sequences that match the query. This allows the user to account for the effects of sampling bias on prevalence estimates. |
| What is the latest research available on this lineage and/or set of mutations? | With the growth of new variants over the pandemic, we have seen many studies that focus on important aspects of a lineage such as the ability to evade immune response and the impact on vaccine efficacy. In order to aid in the discoverability of preprints, publications, datasets and other resources, we show the entries that match a given lineage or mutation query from our up-to-date Research Library <sup>32</sup> (**Fig 2i**). |

### Location Tracker

Some VOCs have only been regionally dominant. For example, Beta and Gamma were dominant in South Africa^8^ and Brazil^35^ respectively. Similarly, B.1.621^36^ was only dominant in Columbia, A.2.5 was only dominant in Panama, and B.1.177 exhibited a high growth rate only in European countries due to a resurgence of travel in the summer of 2020^24,37^. Factors such as the attack rate, population immunity due to previous infection or vaccination, and social mobility vary from one region to the next and have a significant impact on the growth rates exhibited by a given lineage. To account for such localized factors, it is important to have the ability to track the growth of lineages at different geographic scales. We built the Location Tracker on outbreak.info to facilitate the surveillance of SARS-CoV-2 lineages at a country, state/province, or county/city level. The Location Tracker provides a snapshot of circulating lineages with a focus on the last 60 days, and allows users to compare the prevalence of a customizable set of lineages/mutations over time in that location. Furthermore, the tracker also integrates reported cases over time to provide insights on the impact of growth of various lineages on caseloads in the region. As with the Lineage/Mutation Tracker, we designed the user interface to answer a set of specific questions as shown in Table 2.

**Table 2.** Questions addressed by the Location Tracker.

| Question | Relevant visual elements |
| --- | --- |
| What are the most prevalent lineages over the last 60 days? | In order to quickly provide a snapshot of the lineages currently circulating in a given location, we show a stream graph of the prevalence of lineages over the last 60 days (**Fig 3a**). In order to increase interpretability, we grouped lineages that are below 3% prevalence for at least five days over the last 60 days into a separate category, "Other". The prevalence over time can be skewed especially in recent days due to the lag between sample collection, sequencing, and the deposition of sequence data. To convey this uncertainty, the total number of samples collected are shown in an inverted bar graph below the stream graph. In addition, a stacked bar graph shows a snapshot of the cumulative prevalence of the lineages over the last 60 days (**Fig 3b**). Additionally, the user can adjust this window to look at different time windows, e.g. 180 days. |
| What is the distribution of mutations across these lineages? | The Location Tracker shows a snapshot of currently circulating lineages which will help identify a newly emerging lineage that exhibits a high relative growth rate. Often in such cases, the mutations found in the lineage might provide preliminary evidence on phenotypes exhibited by |
|  | the virus such as increased transmissibility or immune evasion. To facilitate this process, we show the prevalence of mutations that are present in the spike gene of at least 75% of the sequences of currently circulating lineages (**Fig 3c**). A Lineage Comparison Tool is also available which expands upon this functionality with customizable queries to add lineages based on the name, VOC/VOI classification, prevalence of mutations, and prevalence within a location. |
| How does the prevalence of different lineages or mutations within this location change over time? | In addition to showing a snapshot of the lineages circulating over the last 60 days, we developed a component to show the temporal variation in the prevalence of a customizable set of lineages/mutations for a given location. This offers additional flexibility to dynamically select lineages or mutations of interest and compare their prevalence over time with a customizable time window (**Fig 3d**). |
| How does the lineage prevalence over time correspond to the number of daily reported cases in this region? | The impact of lineage dynamics on the reported cases over time is of primary concern to public health. To accomplish this, we cross-linked the reported cases for each location using a standardized location identifier, and this is shown in a line graph below the prevalence of a lineage (**Fig 3e**). In addition, users can select a time range within the prevalence chart or the reported cases chart to compare trends over a shorter time span. |

### Case Study: outbreak.info as a hypothesis generation tool to investigate geographic variation in lineage dynamics of VOCs

As the pandemic has continued to progress, we have seen the emergence of VOCs with significant fitness advantages that were able to outcompete previously circulating lineages. As of May 2022, there have been five designated VOCs: Alpha (B.1.1.7 + sublineages, indicated by *), Beta (B.1.351*), Gamma (P.1*), Delta (B.1.617.2*), and Omicron (B.1.1.529*). Of these, Alpha, Beta and Gamma were estimated to have emerged between September and December 2020^9,38,39^ and were subsequently outcompeted globally by the Delta variant that was first detected in December 2020^40^. The Omicron lineage, first detected in November 2021^11^, was able to outcompete Delta and grew much more rapidly relative to previous VOCs during their emergence (**Fig 4a**). Whereas Delta and Omicron variants exhibited high growth rates with little variation globally, Alpha continued to circulate in low prevalence in Brazil and South Africa, where Gamma and Beta variants were dominant respectively (**Fig 4b, 5c**). Additionally, the prevalence of sublineages within Delta and Omicron variants varies geographically. The Location Tracker on outbreak.info can be used to track the growth of VOCs within a given location, thus facilitating the comparison of lineage growth rates across locations. The Location Tracker can also be used to track the relative prevalence of sublineages within these VOCs, shedding light on any geographic variation in these dynamics. Here, we examine trends in the prevalence of the five VOCs globally and highlight the geographic variation in growth rates of Alpha, Beta, Gamma, Delta, and Omicron variants.

**Figure 3.**
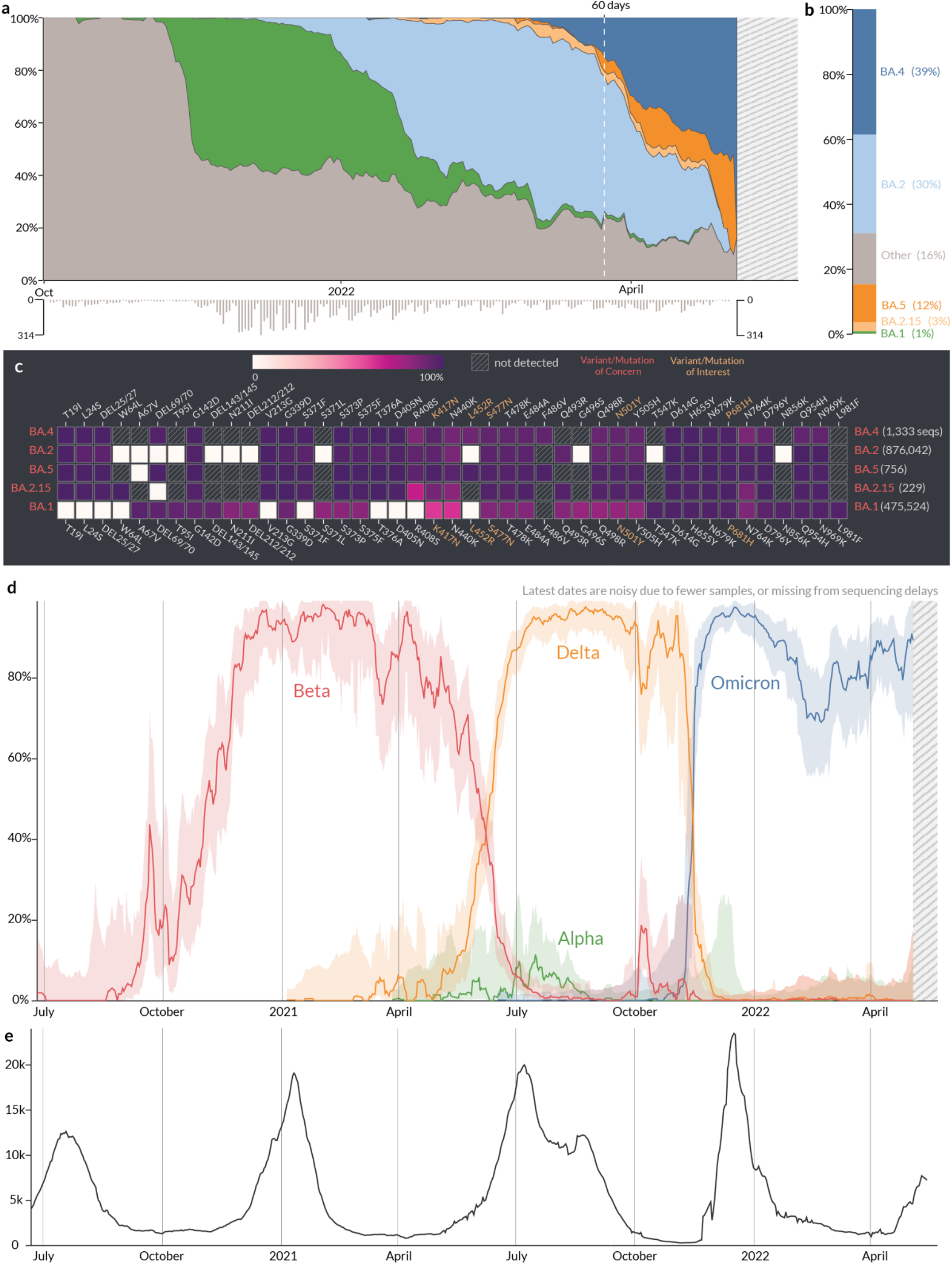
Location report. **a**, Relative prevalence of all lineages over time in South Africa. Total number of sequenced samples collected per day are shown in the bar chart below. **b**, Relative cumulative prevalence of all lineages over the last 60 days in South Africa. **c**, Mutation prevalence across the most prevalent lineages in South Africa over the last 60 days. **d**, Comparison of the prevalence of VOCs grouped by WHO classification: Alpha, Beta, Delta, and Omicron over time in South Africa. **e**, Daily reported cases in South Africa are shown in the line chart below.

**Figure 4.**
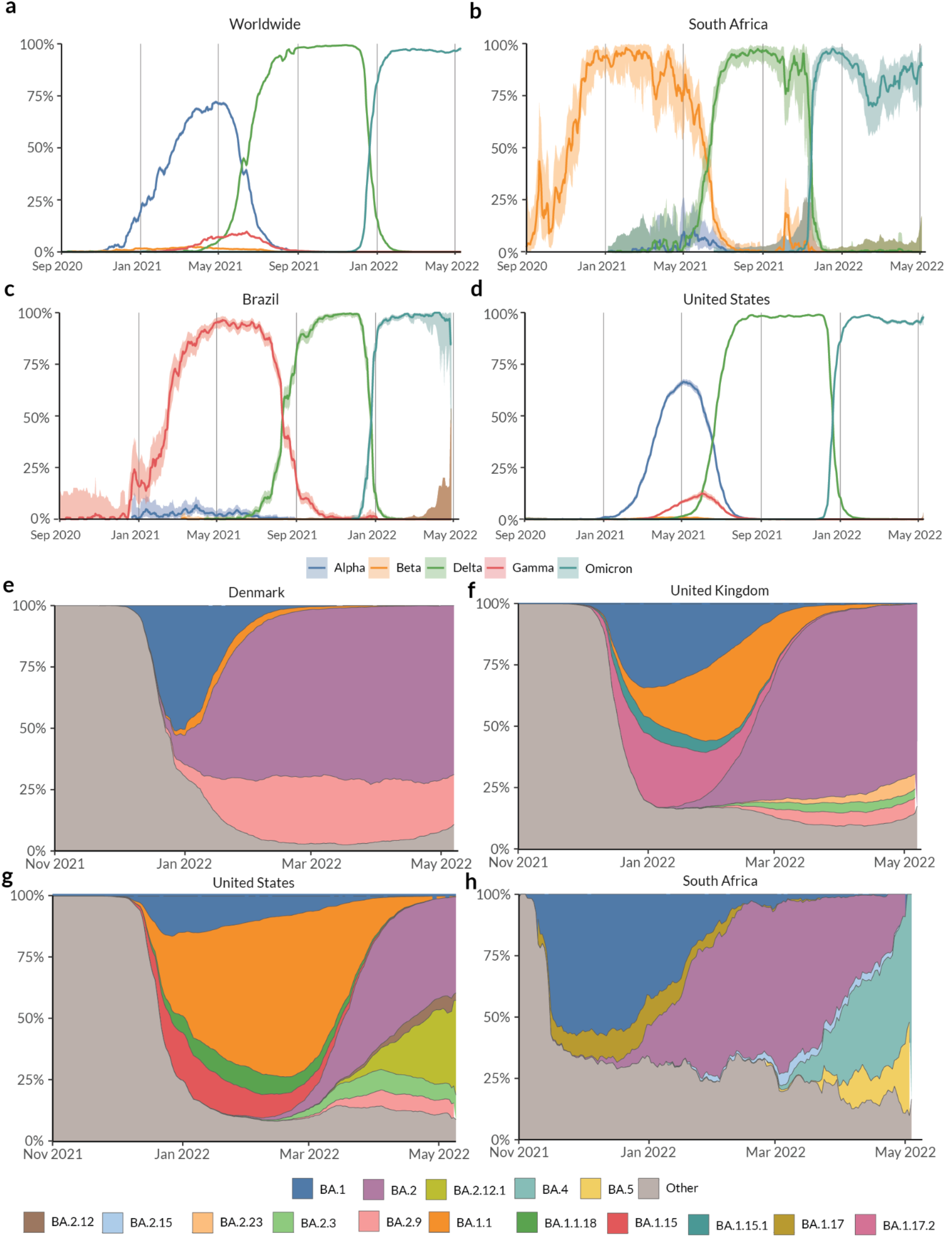
Prevalence of Variants of Concern: Alpha, Beta, Gamma, Delta, and Omicron lineages over time in the **(a)** Worldwide, **(b)** South Africa, **(c)** Brazil, and **(d)** United States. Lineages with a prevalence over 3% over the last 60 days in **(e)** Denmark, **(f)** United Kingdom, **(g)** United States, and **(h)** South Africa.

The earliest samples of the Alpha variant were sequenced in Southern England in late September 2020^38^. There were multiple introductions of the lineage into the United States (U.S.) as early as late November^41^. The Alpha variant showed a transmission advantage of 40-50% in the U.S.^26^, in line with observations in the United Kingdom and the Netherlands. In the U.S., Alpha was able to outcompete previously circulating lineages and continued to increase in prevalence until the introduction of the Delta variant around April 2021 (**Fig 4d**). In contrast to the U.S., the Alpha variant circulated at very low prevalence in Brazil, while the Gamma variant remained dominant in the country^9^ until the introduction of the Delta variant around April 2021 (**Fig 4b**). Similarly, in South Africa, the Beta variant continued to spread until the emergence of the Delta variant and the Alpha variant never became dominant (**Fig 4c**). Whereas the Beta and Gamma variants were able to outcompete Alpha in South Africa and Brazil respectively, Gamma only reached a maximum prevalence of 8% in the U.S. in May 2020, and Beta circulated at a prevalence of <1% (**Fig 4d**). The growth of a lineage is determined by epidemiological factors such as number of introductions, travel between locations, and by intrinsic biological properties such as transmission advantage or immune evasion. Both Beta and Gamma variants show varying degrees of immune evasion^42^. Regions of Brazil had attack rates as high as 75% in October 2020^43^, indicating that immune evasion was the primary reason for the rapid growth of the P.1 lineage in Brazil. In contrast, states in the U.S. had an estimated attack rate between 0.1% and 16% in June 2020^44^. Given this difference in attack rates, we can hypothesize that the intrinsic transmission advantage of the Alpha variant was able to outcompete the advantage conferred by immune evasion of Gamma in the U.S., but the opposite was true in Brazil and South Africa. In all three countries, the introduction of the Delta lineage displaced previously circulating Alpha, Beta, and/or Gamma lineages in the summer of 2021.

The Delta variant of SARS-CoV-2 was first detected in Maharashtra, India in December 2020^40^, has been shown to be 40%-60% more transmissible than Alpha^45,46^, and causes a reduction in vaccine efficacy relative to previously circulating lineages^47^. Vaccination campaigns against COVID-19 started in December 2020 and despite the progress of these campaigns^48^, the Delta variant continued to cause a renewed surge in infections globally. The Delta variant report, which can be accessed directly on the landing page of the lineage reports view, can be used to understand the dynamics of its sublineages. **Fig 4a** shows the global prevalence of the Delta variant over time. This growth reflects the transmission advantage that Delta has over previously circulating lineages including VOCs Alpha, Beta, and Gamma. As the Delta variant continued to spread, its genetic diversity increased and as of May 2022, over 200 sublineages of Delta have been designated^49^.

The Omicron variant was first detected in November 2021 by genomic surveillance teams in South Africa and Botswana. This variant was associated with a rapid resurgence of infections in Gauteng Province, South Africa and was designated a VOC by the WHO within 3 days of uploading the first genome^11^. The variant grew in prevalence very rapidly and within three weeks, the variant was detected in 87 countries and as of May 2022, Omicron has a prevalence of over 95% globally (**Fig 4a**). While increased transmissibility confers a bigger fitness advantage compared to immune evasion when population immunity is low, the opposite is true as population immunity increases either due to vaccination or previous infection^50^. The Omicron variant was found to have a five fold higher chance of reinfection compared to Delta^51^, and Omicron infections presented with a higher viral load than wild type but still lower than Delta^52^. As viral load is one of the determinants of transmissibility, this indicates that Omicron is intrinsically not as transmissible as Delta, but it exhibits better immune evasion. This combination gave Omicron a large fitness advantage over Delta as evidenced by its rapid growth rate worldwide (**Fig 4a**). The continued spread of the variant has resulted in the emergence of many sublineages and as of May 2022, over 100 sublineages of Omicron have been designated. Importantly, there is significant geographic variation in the relative prevalence of newly designated sublineages such as BA.2.12.1, BA.4, and BA.5. While BA.2 continues to be the dominant sublineage within Omicron in countries such as Denmark and the United Kingdom (**Fig 4e, 4f**), we see the BA.2.12.1 sublineage slowly displacing BA.2 in the United States (**Fig 4g**). In South Africa, sublineages BA.4 and BA.5 have completely displaced the previously dominant BA.2 (**Fig 4h**) and have led to another surge in reported cases (**Fig 3e**). The three variants, BA.2.12.1, BA.4, and BA.5 have been shown to evade antibodies elicited by prior BA.1 infection in *in vitro* neutralization studies^53,54^. This observed escape was higher than what was observed for BA.2^55^, highlighting the possibility that these variants led to a renewed surge in infections as these variants continue to spread globally. While the growth of Alpha and Delta variants globally was driven primarily by higher intrinsic transmissibility, the growth of the new variants within Omicron seems to be driven primarily by enhanced immune evasion. The increasing prevalence of immunity due to vaccination or prior infection worldwide, further supports this hypothesis.

This case study illustrates how outbreak.info can be used to not only track and compare the prevalence of lineages across locations, but also to derive and support hypotheses regarding the complex interplay between epidemiology and the intrinsic phenotypic characteristics of emerging SARS-CoV-2 lineages as the virus continues to spread.

## Discussion

The Omicron variant, first detected in late November 2021, has outcompeted Delta and is currently the most dominant lineage globally. However, it is important to note that regardless of how prevalent previously circulating VOCs were, all five VOCs emerged independent of each other. While the current hypothesis for the emergence of VOCs is prolonged virus evolution in a chronically infected individual^56^, we still lack a thorough understanding of this process. Given the underlying stochasticity of this process, *predicting* the emergence of a new VOC is not currently feasible. As a result, continued surveillance of all currently circulating lineages is of utmost importance to public health globally — particularly as SARS-CoV-2 continues to spread and evolve worldwide.

The global community has generated over 10 million genomes of SARS-CoV-2 as of May 2022, shared on platforms such as GISAID^14^. The wealth of primary genomic data can enable downstream applications such as tracking the prevalence of different virus lineages in near real-time. However, the sheer volume of genomic data that continues to increase daily presents challenges to running analyses ad hoc. We developed outbreak.info to serve as a template for tracking the spread of the pandemic over varying geographic and temporal scales at scale, across the world, in near-real time. This new paradigm centralizes the computation of key statistics based on the analysis of disparate data streams. We designed the server infrastructure of outbreak.info keeping two goals in mind: **scalability** of the API as existing data sources increase in size and new data sources are incorporated and **reusability** of the computed data by providing programmatic access through an R package (**Fig 5**). This approach enables us to quickly adapt to and incorporate new modes of surveillance such as the CDC’s National Wastewater Surveillance System^57^. Furthermore, the easy dissemination of any computed data on outbreak.info via the R package enables users to further interrogate and utilize this data for more sophisticated downstream analyses. To maximize accessibility of these data, the web interface of outbreak.info has been designed to offer a high degree of customizability, allowing users to answer specific biological questions and use the platform as a hypothesis generation tool. The guiding principles for the web interface have been **interactivity** via responsive user interface (UI) elements powered by a high performance API, and **interpretability** via intuitive visualization of data based on discussions with researchers, epidemiologists, and public health officials.

**Figure 5.**
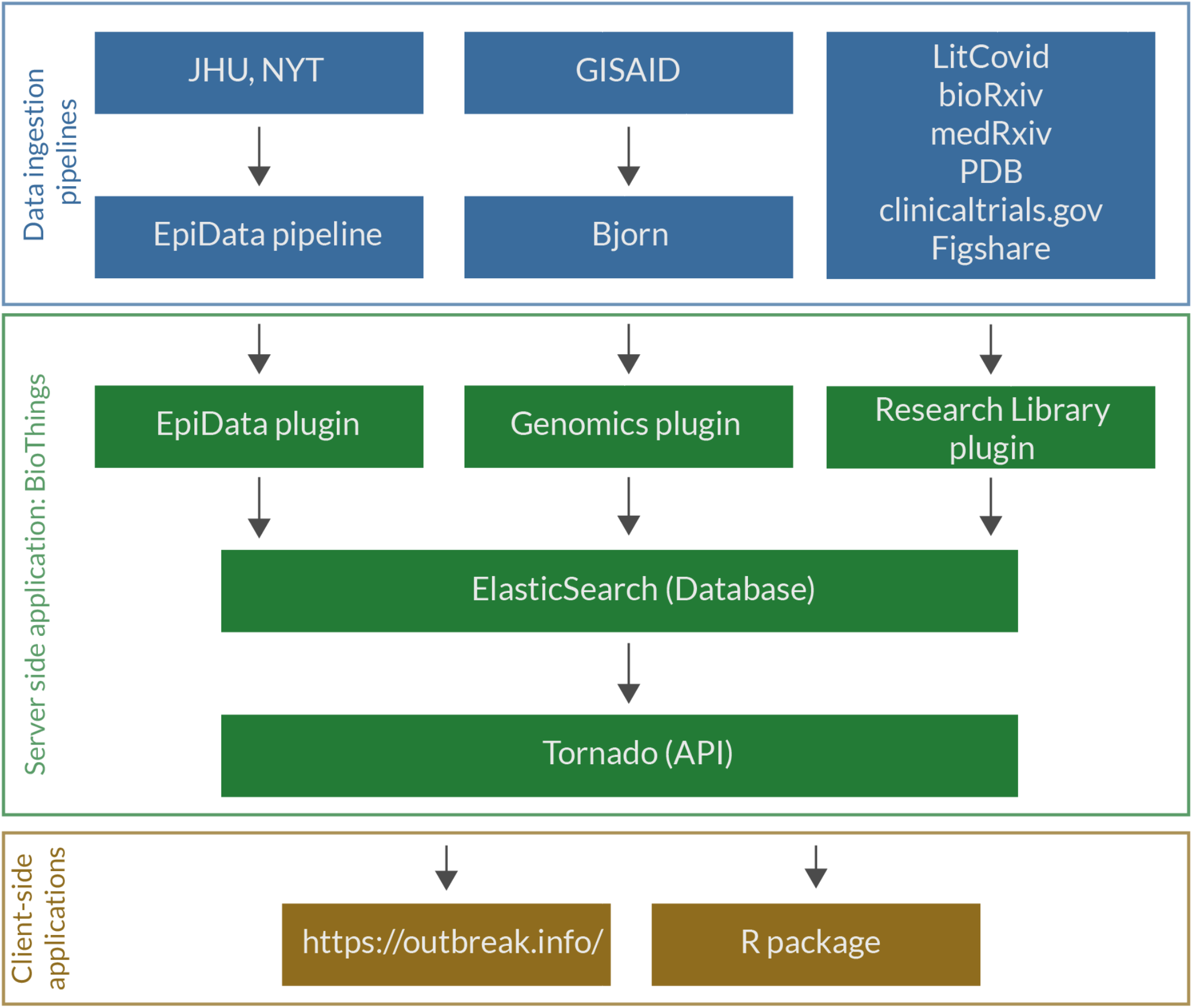
Software infrastructure of outbreak.info. The infrastructure can be broadly divided into (1) Data ingestion pipelines, (2) Server-side hosting the database and API server, and (3) Client-side applications that use the API from the server.

outbreak.info has been enabled by unprecedented global genomic sequencing efforts, and we developed every element of the application to fully leverage this capacity. However, genomic sampling varies globally with the vast majority of sequences coming from high income countries; even within well-sampled regions, there is geographic and temporal variation^13^. To communicate the increased uncertainty due to low sampling, we calculate confidence intervals of estimates wherever applicable, provide histograms of sampling density, and mask data when there are very few data points available. Furthermore, sampling can be selective as samples of the Alpha variant and BA.1 lineage (sublineage of Omicron) show S gene target failure on a widely used qPCR assay. Such sampling biases impact the insights that can be derived from quantities such as the prevalence of a lineage/mutation. We communicate these limitations through a dedicated “caveats” page with warnings regarding the interpretation of data interspersed throughout the interface. outbreak.info continues to provide a mechanism for researchers, epidemiologists, and public health officials to easily track the growth of variants, across any number of locations. The platform, backed by robust infrastructure, allows users to quickly access key statistics for known VOCs, emerging variants, and any combination of mutations without having to run any time-consuming analyses. This allows researchers to focus on data exploration, hypothesis generation and more complex downstream analyses. Beyond the SARS-CoV-2 pandemic, outbreak.info serves as a model for providing ***scalable*** and ***reusable*** metrics to track the spread of any pathogen during an outbreak via interactive and interpretable visualizations.

## Methods

### Ingestion of genomic data

We built a data pipeline, `Bjorn`, to count mutations from a given set of genomes in a scalable manner daily (**Fig 6**). The pipeline consists of the following steps: (1) Download SARS-CoV-2 genomes from the GISAID provision; (2) Divide sequences into chunks of 10,000; (3) Align these sequences using minimap2^58^; (4) Convert the alignment into a FASTA file using gofasta (https://github.com/virus-evolution/gofasta); (5) count mutations and deletions from this alignment; (6) standardize and filter the metadata: country, division, location, pangolin lineage, date of collection, and date of submission and (7) combine results from all chunks and convert to a JSON object. We standardized geographic identifiers using shapefiles from GADM^59^. The final JSON object is loaded into an Elasticsearch index within the BioThings framework^60^. The code for `Bjorn` is available at https://github.com/andersen-lab/bjorn.

**Figure 6.**
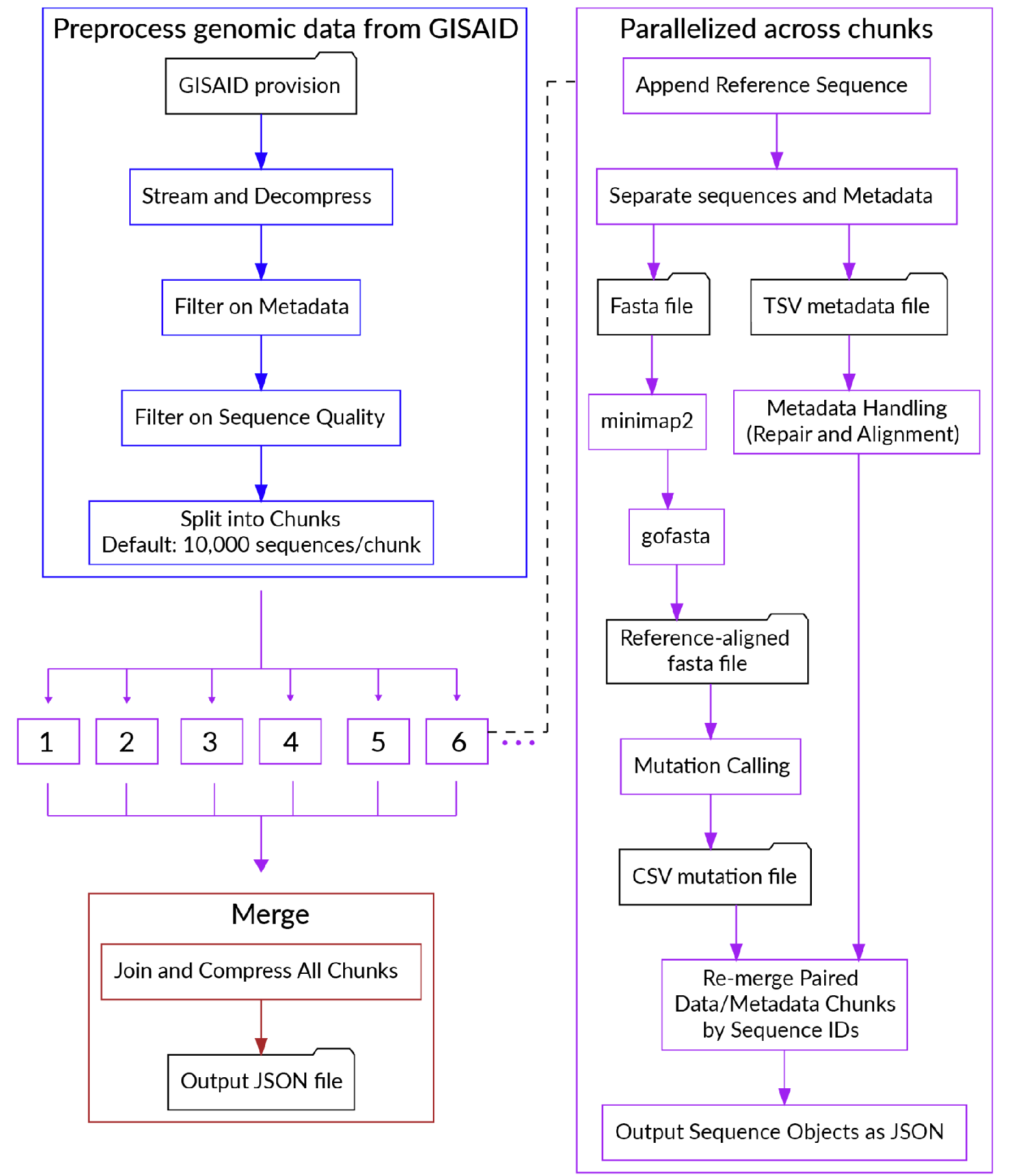
Flowchart describing the steps in `Bjorn`.

### Ingestion of epidemiological data

We built the EpiData pipeline to ingest reported global cases, and deaths from Johns Hopkins University^31^. We used shapefiles from Natural Earth^61^ to standardize geographic identifiers, and obtain populations for countries and states outside the U.S. For the U.S., we used the county level shapefiles and population estimates from the 2019 population estimates by the Census Bureau to standardize geographic identifiers and get population estimates. We standardized reported date formats, and geographic identifiers across the two resources. The code for the EpiData pipeline is available at https://github.com/outbreak-info/biothings_covid19.

### Calculation of confidence intervals on prevalence

Most estimates of prevalence on outbreak.info are binomial proportions. We calculate 95% confidence intervals for these estimates using a Jeffrey’s Interval, the 2.5 and 97.5 quantiles of the beta distribution *β*(*x* + *0*.*5, n* − *x* + *0*.*5*) where *x* is the number of successes and *n* is the number of trials.

### Creation of outbreak.info API

In order to scale with the increasing size of existing data sources and the heterogeneity of newly emerging data sources, we used the BioThings framework^60^. The JSON outputs of our data pipelines are ingested by the BioThings framework and the processed data is stored in individual Elasticsearch indices. A Tornado server is used to create API endpoints that leverage the search capabilities of Elasticsearch to perform complex aggregations of the underlying data. These API endpoints allow the client-side applications to query the underlying data within reasonable query times while accounting for the scale of the ingested data. The BioThings Hub maintains historical data by default, allowing us to roll back to previous data backups if issues are discovered with new data after they are deployed. The code for the server-side application is available at https://github.com/outbreak-info/outbreak.api.

### outbreak.info web application

The web application was built using Vue.js^62^, a model–view–viewmodel JavaScript framework which enables the two-way binding of user interface elements and the underlying data allowing the user interface to reflect any changes in underlying data and vice versa. The client-side application uses the high performance API to interactively perform operations on the database. Customized data visualizations on the client were built using D3.js^63^, giving us the ability to develop novel, and intuitive visual elements as part of the user interface. We designed these visualizations to answer specific questions of interest to epidemiologists, researchers, and public health officials. We further added functionality to enable the 1-click copy or download of every chart in the interface as a png or svg. The code for the client-side application is available at: https://github.com/outbreak-info/outbreak.info

### R package

We developed an R package for outbreak.info to allow researchers and other individuals to easily access the data via the API for downstream analyses and visualizations. The R package is composed of three parts: functions that allow the user to access genomic data, functions to access the epidemiological data, and functions to access the Research Library metadata. They all consist of a base function that contains arguments for all possible parameters that can be used to query the API. While users can utilize this base function directly to access data, several wrapper functions are available that inherit the arguments from the base function in addition to pre-specified parameters to simplify the process of querying the API. For example, while `getGenomicData()` can be used directly to access data regarding the daily global prevalence of a specified lineage, doing so would require a user to be familiar with the name of the endpoint as specified in the API URL (in this case, global-prevalence). Instead, the user can access this data with the more intuitively named `getPrevalence()`.Therefore, these wrapper functions allow users to easily and quickly obtain the data they need. The R package also contains an authenticateUser() function that allows users to authenticate against their GISAID credentials and access computed statistics from the primary genomic data provided by GISAID.

In addition, as the API queries location by ISO3 code, rather than by location name, two functions have been created that allow users to forgo the step of searching for the ISO3 code themselves: `getISO3Code()` and `getLocationIdGenomic()`. The latter function uses the genomics API endpoint to obtain the ISO3 code for a given location. The ISO3 code can be obtained with either a full or incomplete location name; in the latter case, the user will be provided a list of matching locations and must specify the location they are interested in. This function is embedded in the parent `getGenomicData()` function, and is therefore inherited in all wrapper functions. Therefore, searching for data by location in the R package replicates the experience on the client-side web application. Documentation is available at: https://outbreak-info.github.io/R-outbreak-info with vignettes located at https://outbreak-info.github.io/R-outbreak-info/articles/index.html. The R package can be downloaded and installed using the remotes package function: `install_github(“outbreak-info/R-outbreak-info”)`.

## Data Availability

All data produced are available online at https://outbreak.info/

## Acknowledgements

We thank the technical team, sequence curators, and administrators of the GISAID database for helping us with this project and supporting rapid and transparent sharing of genomic data during the COVID-19 pandemic. We thank our colleagues for sharing genomic data on GISAID. This work was supported by the National Institute for Allergy and Infectious Diseases (5 U19 AI135995, 3 U19 AI135995-04S3, 3 U19 AI135995-03S2, U01AI151812, R01 AI162611, R01 AI153044), National Center For Advancing Translational Sciences (5 U24 TR002306, UL1TR002550), Centers for Disease Control and Prevention (75D30120C09795), and the National Institute of General Medical Sciences (R01GM083924).

