## Supplemental File 1 for "Outbreak.info genomic reports: scalable and dynamic surveillance of SARS-CoV-2 variants and mutations"

Clement Png Wen Jie[1,2]
Constanza Schiavina[1]
Felipe André Silva[1]
Gabriela Calegario[1,3]
Giovanni Marques de Castro[1]
Joses Ho[1,2]
Juan Finello[1]
Letícia Maria Rodrigues[1]
Lucas Freitas[1,3]
Meera Makheja[1,2]
Mikhail Bakaev[1,5]
Motharasan Manogaran[1,4]
Paola C Resende[1,3]
Priscila Born[1,3]
Shruti Khare[1,2]
Sofia Romano[1]
Suma Tiruvayipati[1,6]
Swathi Nachiar Manivannan[1,2]
Tze-Minn Mak[1,2]
Ya Ni Xu[1,2]
Yi Hong Chew[1,2]

[1]GISAID Global Data Science Initiative (GISAID), Munich, Germany,
[2]Bioinformatics Institute, Agency for Science Technology and Research, Singapore,
[3]Oswaldo Cruz Foundation (FIOCRUZ), Rio de Janeiro, Brazil,
[4]National Institutes of Biotechnology Malaysia, Selangor, Malaysia,
[5]Smorodintsev Research Institute of Influenza, St. Petersburg, Russia,
[6]Genome Institute of Singapore, Agency for Science Technology and Research, Singapore
